# High efficacy of saliva in detecting SARS-CoV-2 by RT-PCR in adults and children

**DOI:** 10.1101/2020.12.01.20241778

**Authors:** Michael Huber, Peter W. Schreiber, Thomas Scheier, Annette Audigé, Roberto Buonomano, Alain Rudiger, Dominique L. Braun, Gerhard Eich, Dagmar I. Keller, Barbara Hasse, Jürg Böni, Christoph Berger, Huldrych F. Günthard, Amapola Manrique, Alexandra Trkola

**Affiliations:** Institute of Medical Virology, University of Zurich, Zurich, Switzerland; Division of Infectious Diseases and Hospital Epidemiology, University Hospital Zurich and University of Zurich, Zurich, Switzerland; Division of Infectious Diseases and Hospital Hygiene, Spital Limmattal, Schlieren, Switzerland; Division of Medicine, Spital Limmattal, Schlieren, Switzerland; Division of Infectious Diseases, Hospital Hygiene and Occupational Medicine, Stadtspital Triemli, Zurich, Switzerland; Emergency Department, University Hospital Zurich, Zurich, Switzerland; Division of Infectious Diseases and Hospital Epidemiology, University Children’s Hospital Zurich, Zurich, Switzerland

**Keywords:** SARS-CoV-2, PCR, pediatric, saliva

## Abstract

**Background:** RT-PCR of nasopharyngeal swabs (NPS) is the acknowledged gold standard for the detection of SARS-CoV-2 infection. Rising demands for repetitive screens and mass-testing necessitate, however, the development of additional test strategies. Saliva may serve as an alternative to NPS as its collection is simple, non-invasive and amenable for mass- and home-testing but rigorous validation of saliva particularly in children is missing.

**Methods:** We conducted a large-scale head-to-head comparison of SARS-CoV-2 detection by RT-PCR in saliva and nasopharyngeal swab (NPS) of 1270 adults and children reporting to outpatient test centers and an emergency unit for an initial SARS-CoV-2 screen. The saliva collection strategy developed utilizes common, low-cost plastic tubes, does not create biohazard waste at collection and was tailored for self-collection and suitability for children.

**Results:** In total, 273 individuals were tested SARS-CoV-2 positive in either NPS or saliva. SARS-CoV-2 RT-PCR results in the two specimens showed a high agreement (Overall Percent Agreement = 97.8%). Despite lower viral loads in saliva of both adults and children, detection of SARS-CoV-2 in saliva compared well to NPS (Positive Percent Agreement = 92.5%). Importantly, in children, SARS-CoV-2 infections were more often detected in saliva than NPS (Positive Predictive Value = 84.8%), underlining that NPS sampling in children can be challenging.

**Conclusions:** The comprehensive parallel analysis reported here establishes saliva as a generally reliable specimen for the detection of SARS-CoV-2 with particular advantages for testing children that is readily applicable to increase and facilitate repetitive and mass-testing in adults and children.

**Article Summary Main Points:** Comparison with nasopharyngeal swabs in a large test center-based study confirms that saliva is a reliable and convenient material for the detection of SARS-CoV-2 by RT-PCR in adults and increases detection efficacy in children.

## Introduction

The current gold standard for the diagnosis of Severe Acute Respiratory Syndrome Coronavirus-2 (SARS-CoV-2) infection relies on the detection by quantitative reverse-transcription polymerase chain reaction (RT-qPCR) in nasopharyngeal swabs. A range of RT-qPCRs methods have been developed and proven highly sensitive, accurate and reliable [1, 2]. Nasopharyngeal swabs (NPS) are considered the optimal material for detection, particularly in early infection [2]. However, viral load in the nasopharynx can wane in later disease stages, while the virus remains detectable in alternate specimen such as bronchoalveolar lavage or sputum, necessitating a validation of diagnostics tests in these specimens [3-5]. In addition, to overcome limitations in mass screening for early detection of SARS-CoV-2, saliva has been considered as alternate material to NPS [6-10]. NPS collection requires trained personnel while saliva collection is comparatively easy, needs little instruction and is amenable for self-collection. Importantly, saliva collection is non-invasive, and it does not create discomfort for the patient. Saliva would thus be of particular advantage for testing children, for whom often parents and pediatricians refrain from testing due to the need to conduct a nasopharyngeal swab. Likewise, the possibility to switch to saliva would also be a relief for adults when frequent testing or large-scale screens are required, respectively. Further, considering the current high level of SARS-CoV-2 testing by RT-PCR and antigen tests, which both require nasopharyngeal swabs, shortage in swab supplies may occur. Establishing the possibility to switch to saliva collection in this situation to allow RT-PCR testing to continue is thus highly advisable.

Several recent studies have evaluated saliva as alternate specimen [6-29]. While these studies generally agree that detection of SARS-CoV-2 in saliva is possible, comparative analyses came to different conclusions, with some studies noting a better performance of saliva, while others found a substantially lower sensitivity. With few exceptions, patient cohorts tested thus far were in most studies relatively small and often included both hospitalized individuals with advanced SARS-CoV-2 infection as well as outpatients who were newly screened for infection, leaving uncertainty in which situation saliva may be best used. The overall sensitivity and thus utility of saliva in comparison to NPS remains thus differentially debated and needs to be defined. To resolve these issues, we embarked on a large-scale head-to-head comparison of saliva and NPS in a test center setting for adults and children. The high number of individuals tested (N = 1270) and the high number of positives detected (N = 273), paired with a true-to-life screening in test centers, empowered a highly controlled analysis which supports the applicability of saliva in routine testing and particularly provides better opportunities for testing children.

## Materials and Methods

### Study population

Adults (N =1100) and children (N = 170) opting for a SARS-CoV-2 test at one of five participating test centers were included. Four centers were dedicated test centers for outpatients (N=3 for adults; N=1 for children) and one was an emergency department. The study population comprised individuals with SARS-CoV-2 related symptoms based on Swiss testing criteria and asymptomatic individuals with relevant exposure to a SARS-CoV-2 index case. Hospitalized patients were not included. Individuals were included without further selection to avoid skewing. Information on symptomatic or asymptomatic status was collected as part of the regular procedure for SARS-COV-2 testing and reporting based on self-evaluation (asymptomatic/mild/strong) by the participants, as they did not see a physician in the test center setting.

### Ethical approval

The Zurich Cantonal Ethics Commission waived the necessity for a formal ethical evaluation based on the Swiss Federal Human Research Act, as the collection of saliva in parallel to a scheduled nasopharyngeal swab induces no risk (Req-2020-00398). No additional personal data beyond the usual information on symptoms and duration required by the FOPH for all SARS-CoV-2 tests in Switzerland was collected. Due to the ethics waiver no informed consent had to be collected.

### Sample collection

Test centers were advised to use their regular swab and virus transport medium (VTM)/universal transport medium (UTM) for nasopharyngeal sampling. Transport media used by the centers included Cobas PCR Medium (Roche), Liquid amies preservation medium (Copan), Virus Preservative Medium (Improviral), and in-house VTM (HEPES, DMEM, FCS, antibiotics, antimycotics).

Collection kits for saliva were supplied to the test centers: one tube for saliva collection (Sarsted 62.555.001) and a separate tube with 3 ml VTM (Axonlab AL0607). The procedure for saliva collection was described in an instruction leaflet (Figure 1). In Study Arm 1, “Basic”, individuals were asked to clear the throat thoroughly and collect saliva one or two times into the same tube (N = 835). As a guidance for the volume of saliva to be sampled, participants were instructed by study teams to collect 0.5 – 1 ml (approx. a teaspoon full). To investigate a possible influence on SARS-CoV-2 detection in saliva through differences in saliva collection, a subset of patients (N = 435) in Study Arm 2, “Enhanced”, was asked to three times clear their throat and collect saliva into the same tube. Emphasis in this study arm was on enhanced throat clearing to ascertain sampling material from the posterior oropharynx. Immediately after saliva collection, VTM was added to the crude saliva and the content mixed through gentle tilting. Saliva was collected directly after NPS and both specimens immediately sent for SARS-CoV-2 RT-PCR testing.

**Figure 1:**
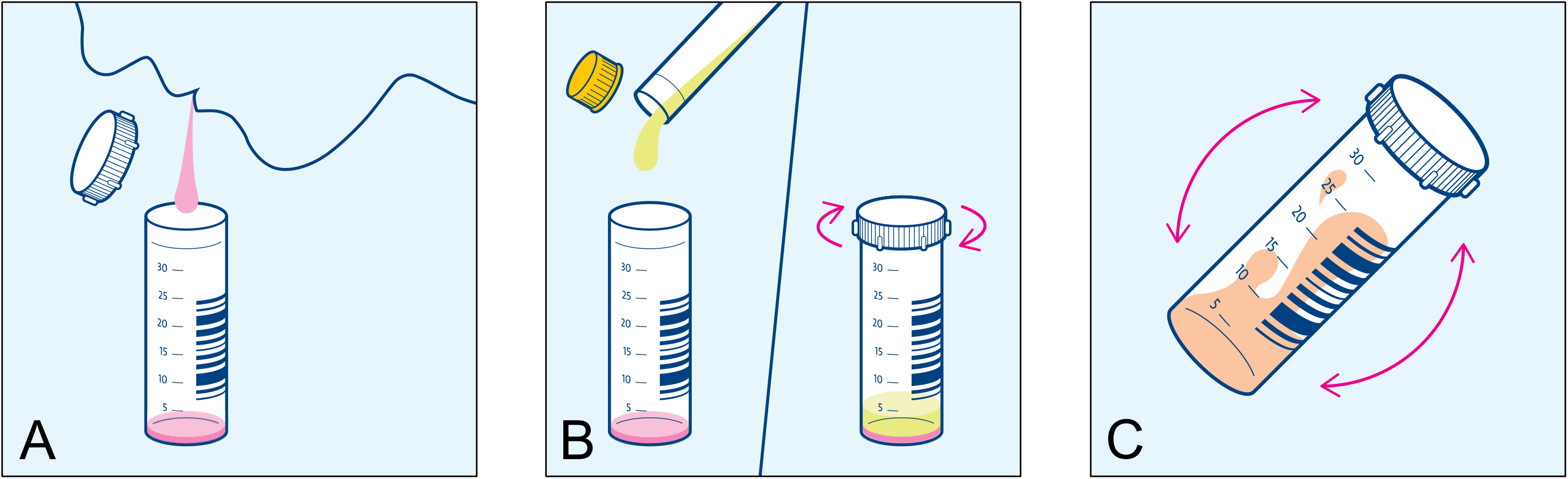
Instruction for saliva collection. Participants were asked to clear the throat and collect saliva into a collection tube (A). VTM was added to the crude saliva immediately after collection (B), and the content was mixed through gentle tilting (C).

### Quantitative SARS-CoV-2 PCR

NPS and Saliva were processed identically using the procedures established for NPS in the diagnostics laboratory of the Institute of Medical Virology. 500 ul of NPS or saliva in VTM were diluted in 500 ul of Nuclisens easyMAG Lysis Buffer (BioMérieux), centrifuged (2000 rpm, 5 min) and analyzed with the Cobas SARS-CoV-2 IVD test (Roche) on a Cobas 6800. All testing for NPS and saliva was done in parallel on the same day. SARS-CoV-2 detection was further quantified using SARS-CoV-2 Frankfurt 1 RNA as calibrator (European Virus Archive, 004N-02005) allowing to report both Ct and genome equivalents.

### Verification by in-house SARS-CoV-2 E-gene and GAPDH PCR

Discordant results of the Cobas SARS-CoV-2 test between NPS and saliva were re-analyzed using an in-house RT-qPCR targeting the E-gen based on Corman et al. [1]. GAPDH was measured as input control as described [30]. Both assays used AgPath-ID One-Step RT-PCR chemistry (Ambion, ThermoFisher).

### Data analysis

E-gene Ct values were used for comparison. If E-gene reported negative but ORF1 reported positive by the Cobas SARS-CoV-2 IVD test, the ORF1 result was considered and the respective sample rated positive for SARS-CoV-2. This was the case for one saliva sample. Data was analyzed using R (version 4.0.2) [31]. 95% confidence intervals were calculated with the epiR package (version 1.0.15). Method comparison and regression analysis (Passing-Bablok Regression [32] and Bland-Altman Plot [33]) was performed with the mcr package (version 1.2.1).

## Results

### Head-to-head comparison of saliva and nasopharyngeal swabs as material for SARS-CoV-2 detection by RT-PCR

We sought to design a saliva collection procedure that is safe, does not create infectious waste at the collection site and does not need trained medical personnel. In order to avoid that contaminated material needs to be disposed at the collection sites, we refrained from using collection tubes with disposable funnels and opted for collection into commonly available, cheap, wide plastic tubes. Our protocol for saliva collection instructs participants to self-collect approx. 0.5 ml saliva into an empty wide (30 ml, 30 mm diameter) tube (Figure 1). Initial attempts in a pilot experiment at the participating emergency department with smaller tubes (15 ml, 17 mm diameter) showed that spitting into narrower tubes is problematic for some participants, leading to a contamination of the outside of the tube with saliva in some cases. Sampling with the wider tubes was in contrast unproblematic and thus deemed safe. Saliva sampling in children was found equally unproblematic; children were collaborating and able to expectorate. We deliberately chose to add VTM after saliva collection and not to use VTM-filled tubes as some persons feel more comfortable with sampling into empty containers. This also circumvents that the VTM is mistakenly used to gurgle or swallowed, issues that have to be considered in home-testing and with children.

Our study included five different test sites to ensure that data are not skewed due to specific procedures at one site. In Study Arm “Basic” (N = 835) saliva sampling was done with one-time throat clearing followed by expectorating saliva one to two times. In Study Arm “Enhanced” (N = 435) sampling was intensified to three times throat clearing and spitting. Saliva was mixed with VTM immediately after collection and shipped to the lab at room temperature the same day or stored at 4°C until shipping the next day.

The thus diluted material was unproblematic for further processing in the laboratory, no complications in pipetting and only rarely samples were rejected due to the viscosity of saliva or invalid results were observed.

### High positive agreement of SARS-CoV-2 detection in saliva and nasopharyngeal swabs

Adults and children that qualified for a regular SARS-CoV-2 test according to the FOPH and reported to one of the participating test centers or emergency units were enrolled from October 20, 2020 to January 28, 2021. In total 1270 individuals (male 54.6%/female 45.4%) were included (Table 1). Median age was 34 with an age range of 5 – 98 years. 170 participants were under the age of 18. The majority of participants were symptomatic 75.6%. Days of symptoms ranged from 1 to 30 with a median of 2 days. The SARS-CoV-2 positivity rate amongst study participants was 21.5%. Gender distribution in children and adults was similar (Table 1). Percentage of symptomatic individuals and participants in the Enhanced study arm was higher in children.

**Table 1:** Participant demographics.

|  | Total (n=1270) | Adults (n=1100) | Children (n=170) |
| --- | --- | --- | --- |
| **Male /** | 693 (54.6%) / | 605 (55%) / | 88 (51.8%) / |
| **Female (%)** | 577 (45.4%) | 495 (45%) | 82 (48.2%) |
| **Age median (range)** | 34 (5 – 98) | 37 (18 - 98) | 13 (5-17) |
| **Symptomatic mild (%)** | 836 (65.8%) | 701 (63.7%) | 135 (79.4%) |
| **Symptomatic strong (%)** | 91 (7.2%) | 79 (7.2%) | 12 (7.1%) |
| **Asymptomatic (%)** | 299 (23.5%) | 279 (25.4%) | 20 (11.8%) |
| **No information on symptoms (%)** | 44 (3.5%) | 41 (3.7%) | 3 (1.76%) |
| **Median days of symptoms (range)** | 2 (1 – 30) | 2 (1 – 30) | 2 (1 – 21) |
| **"Basic" /** | 835 (65.7%) / | 783 (71.2%) / | 52 (30.6%) / |
| **"Enhanced" study arm** | 435 (34.3%) | 317 (28.8%) | 118 (69.4%) |

Across the entire cohort and both study arms NPS and saliva results showed a high overall percent agreement (OPA = 97.8%) and good positive percent agreement (PPA = 92.5%, Figure 2B). In only 28 cases discordant results were observed, with 20 saliva samples and 8 NPS showing a negative result when the other specimen tested positive (Figure 2A). To investigate if discordant results are due to inadequate sampling, detection problems in the RT-PCR, or reflect true negatives in the respective sample material, all discordant pairs were retested using an in-house RT-PCR for the E-gene in conjunction with a GAPDH measurement to control for input. Mean levels for GAPDH input were Ct = 24.3 (SD = 2.6) for NPS and Ct = 24.7 (SD = 2.1) for saliva. One false-negative saliva sample (E-gene Ct 19.7 in NPS) did not contain any material (GAPDH Ct > 40). Excluding this sample, the PPA in the NPS Ct 15 – 20 range reaches 100% (Table 2).

**Table 2:** Positive Percent Agreement (PPA) stratified by NPS E-gene Ct-values.

| NPS (Ct) | >10-15 | >15-20 | >20-25 | >25-30 | >30-33 | >33-35 | >35-40 |
| --- | --- | --- | --- | --- | --- | --- | --- |
| **NPS positive** | 1 | 59 | 96 | 60 | 15 | 13 | 21 |
| **Saliva false negative** | 0 | 1<br>(0*) | 2 | 2 | 0 | 5 | 10 |
| **PPA** | 100% | 98.3%<br>(100%*) | 97.9% | 96.7% | 100% | 61.5% | 52.4% |

\*Excluding one sample that did not contain saliva as defined by GAPDH measurement.

**Figure 2:**
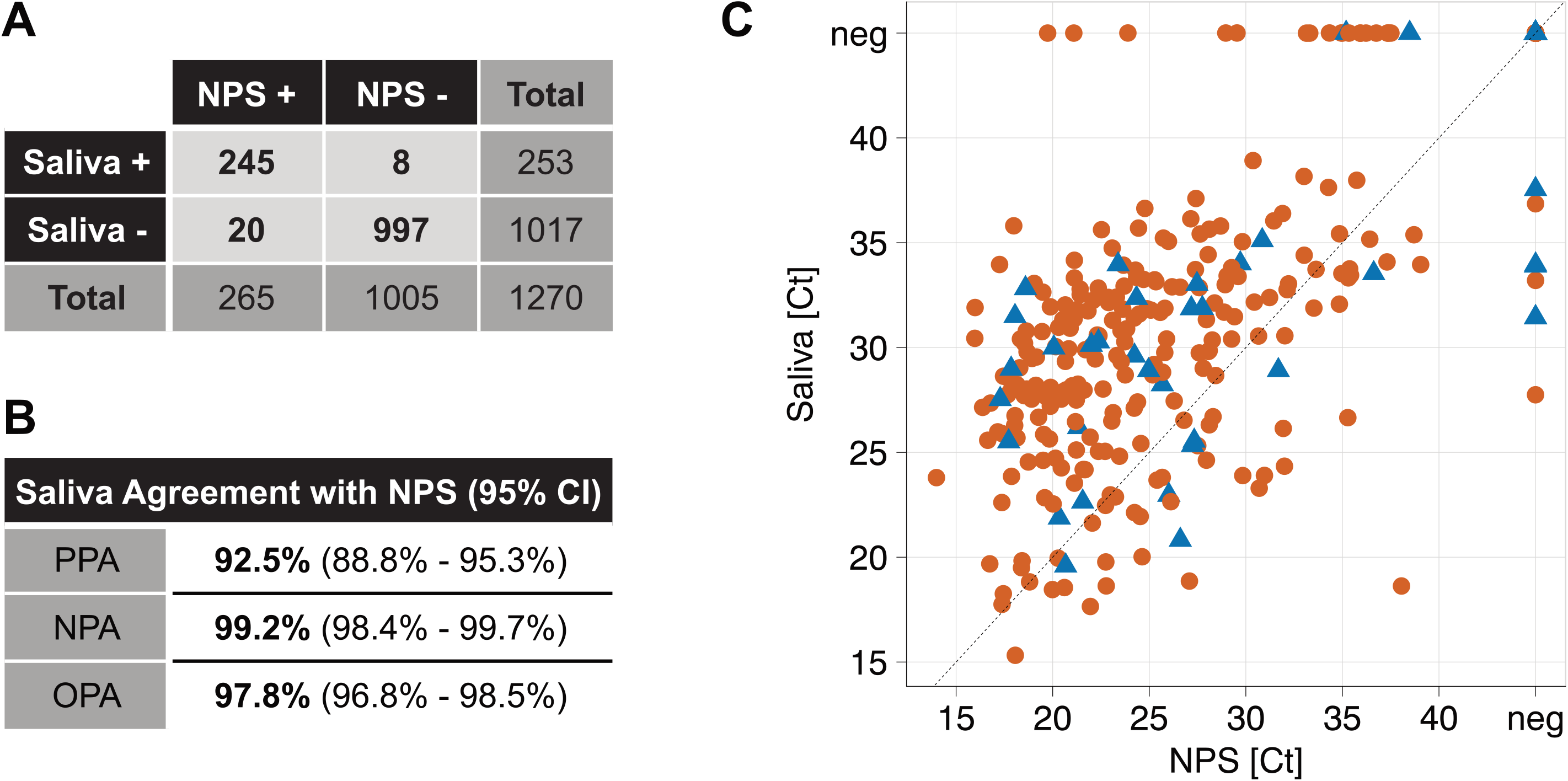
High agreement of SARS-CoV-2 detection in saliva and nasopharyngeal swabs. A) Contingency Table full cohort. B) Agreement values. PPA = Positive Percent Agreement, NPA = Negative Percent Agreement, OPA = Overall Percent Agreement. C) Summary of the full cohort (N = 1270 study participants). Roche Cobas E-Gene Ct values of paired NPS and saliva samples are depicted. Red dots = adults, blue triangles = children, neg = PCR negative, black dashed line equals identity.

Re-assessment with an in-house E-gene PCR confirmed all discordant results. For one case with a negative NPS, a second swab was collected the following day. This sample showed a high viral load, confirming an unsuccessful swab collection the day earlier.

Of note, in our head-to-head comparison both NPS (N = 3) and saliva (N = 5; N = 4 excluding the sample that did not contain saliva) produced false-negative results in cases where the other specimen showed a high viral load (Ct < 33) highlighting variability in collection for both specimens.

Breakdown in adults (Fig 2B) and children (Fig 2C) underline the suitability of saliva for both age groups. Of particular note, children showed more NPS negatives with paired positive saliva (N = 5) than saliva negative/NPS positive pairs (N = 2).

### SARS-CoV-2 loads in saliva and nasopharyngeal swab correlate

Correlation analysis of sample pairs that both tested positive (N = 245) confirmed that saliva and NPS results are in good agreement (Figure 3A_C). Notably, Ct values in saliva were on average 4.87 higher than the corresponding Ct in NPS across the full cohort. Average Ct value differences for NPS and saliva in adults and children were similar (4.93 and 4.44, respectively, Figure 3D). Higher Ct values in saliva correspond to a factor of 29, 30 and 22-times lower viral loads in saliva for the full cohort, adults and children, respectively (Figure 3B). Of note, at high Ct values in the corresponding NPS, the reduction in viral load in saliva was less pronounced possibly adding to the high rate of detection in saliva.

**Figure 3:**
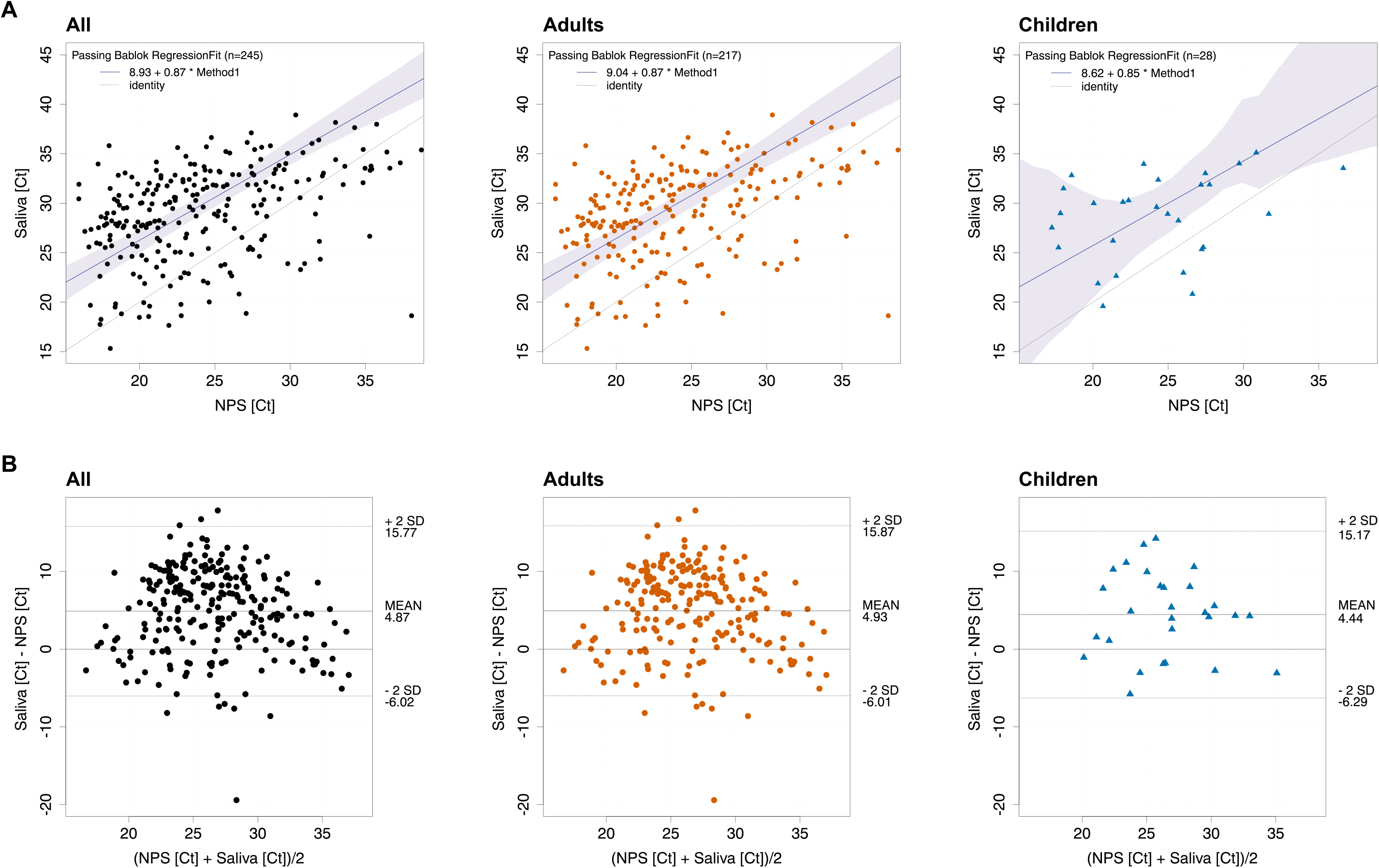
SARS-CoV-2 levels in saliva and nasopharyngeal swabs correlate. A) Passing-Bablok Regression of E-gene Ct-values of NPS and saliva of all positive pairs from the full cohort (N = 245; p < 0.0001), adults (N = 217; p < 0.0001) and children (N = 28; p = 0.079). Black dashed line equals identity, blue line shows linear trend. B) Bland-Altmann Plot of E-gene Ct-values of NPS and saliva of all positive pairs from the full cohort (N = 245), adults (N = 217) and children (N = 28).

### Detection of SARS-CoV-2 in saliva from symptomatic and asymptomatic individuals

Our study recorded severity of symptoms (asymptomatic/mild/strong) at the sampling time point by self-evaluation (Figure 4A). We observed a good positive percent agreement of saliva and NPS in symptomatic individuals in the full cohort (PPA = 92.2%), adults (PPA = 92.9%) and children (PPA = 92.9%). In line with a trend to lower viral loads, i.e. higher Ct values in absence of symptoms (asymptomatic median Ct 28.7; mild symptoms median Ct 23.5; strong symptoms median Ct 21.6), the PPA was lower in asymptomatic participants (PPA = 85.0%). We observed decreasing viral loads with ongoing symptomatic infection in NPS, highlighting a transient window of detection in the upper respiratory tract. Interestingly, changes in saliva were overall less dynamic than in NPS (Figure 4B).

**Figure 4:**
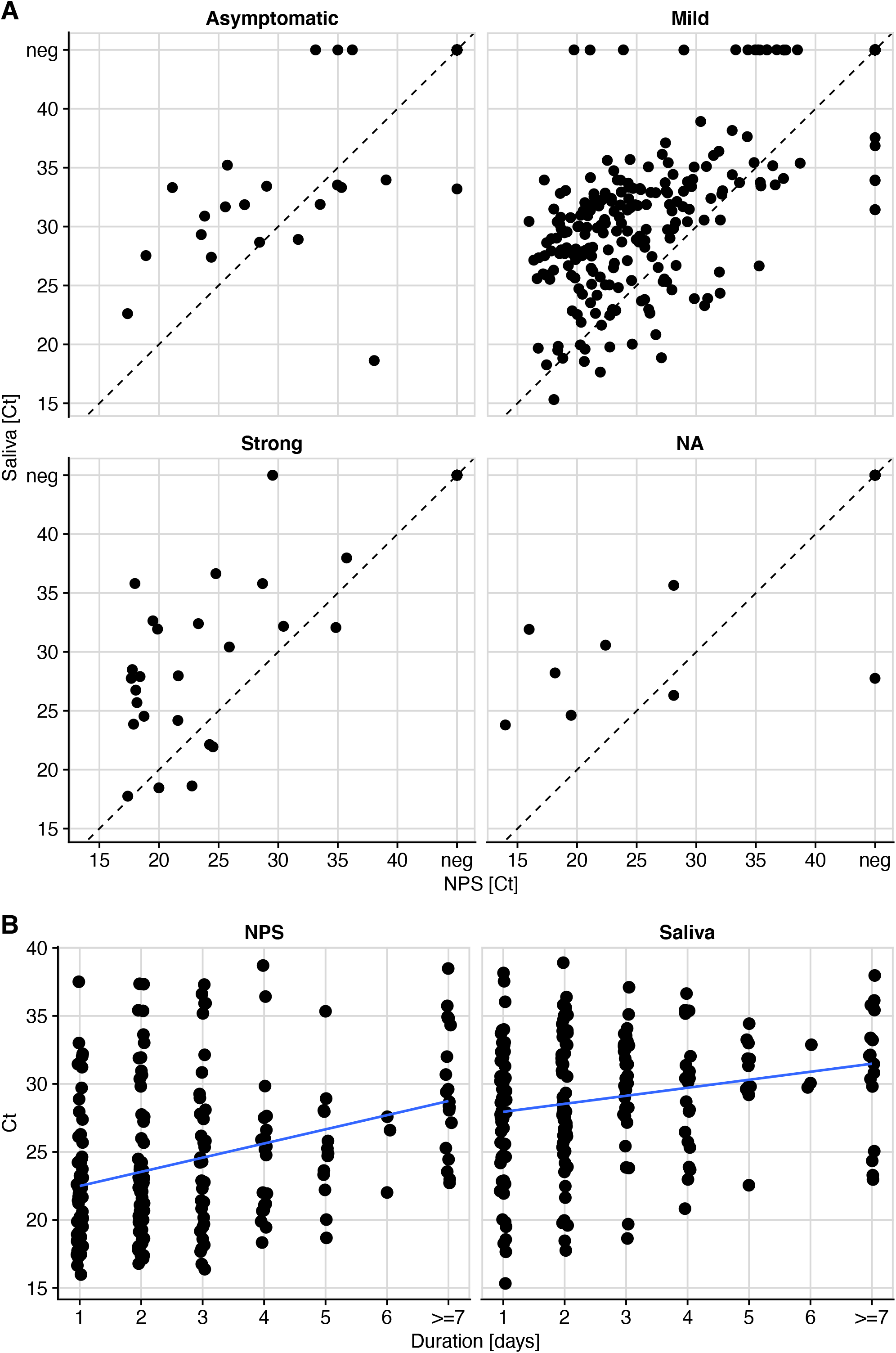
Viral loads in NPS and saliva decrease with ongoing infection. A) E-gene Ct-values of NPS and saliva of all pairs from the full cohort stratified by symptoms. B) Duration of symptoms in symptomatic patients (N = 927) versus E-gene Ct value in saliva and NPS. neg = PCR negative, line shows linear trend.

### Intensified throat clearing with saliva collection is favorable

To investigate if the intensity of saliva collection has an impact, we analyzed the two study arms of saliva collection separately. Participants were either asked to clear the throat thoroughly and collect about 0.5 – 1 ml of saliva (“Basic”, N = 835) or in an intensified protocol to repeat throat clearing and spitting three times (“Enhanced”, N = 435). To ensure appropriate statistical power we investigated the influence of sampling solely in the full cohort. We found that intensified saliva collection appears favorable for samples with low viral load. With the enhanced sampling protocol, PPA with NPS of ct >33 reached 66.7% (CI 35% - 90%), compared to 50.0% (CI 28% - 72%) with the basic protocol (Figure 5 and Table 3). Differences were, however, not significant, highlighting robust detection of SARS-CoV-2 in saliva in the two collection procedures tested.

**Table 3:**
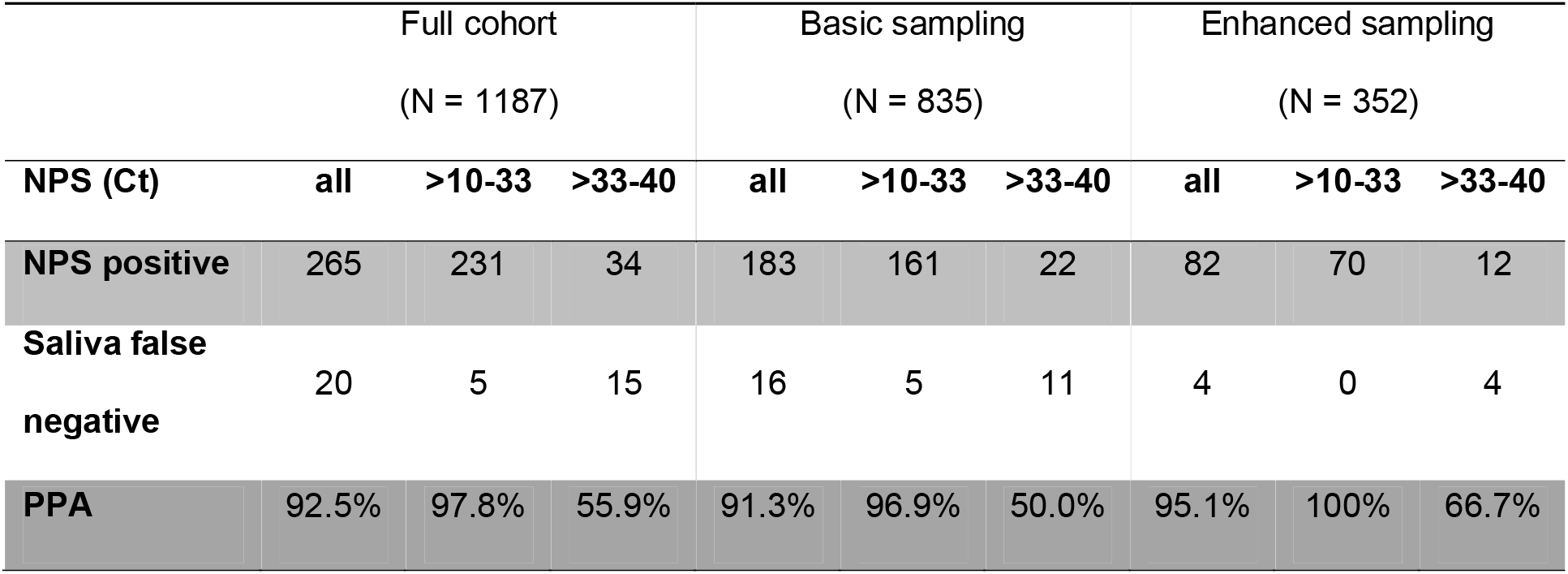
Positive Percent Agreement (PPA) stratified by NPS E-gene Ct-values and saliva sampling.

**Figure 5:**
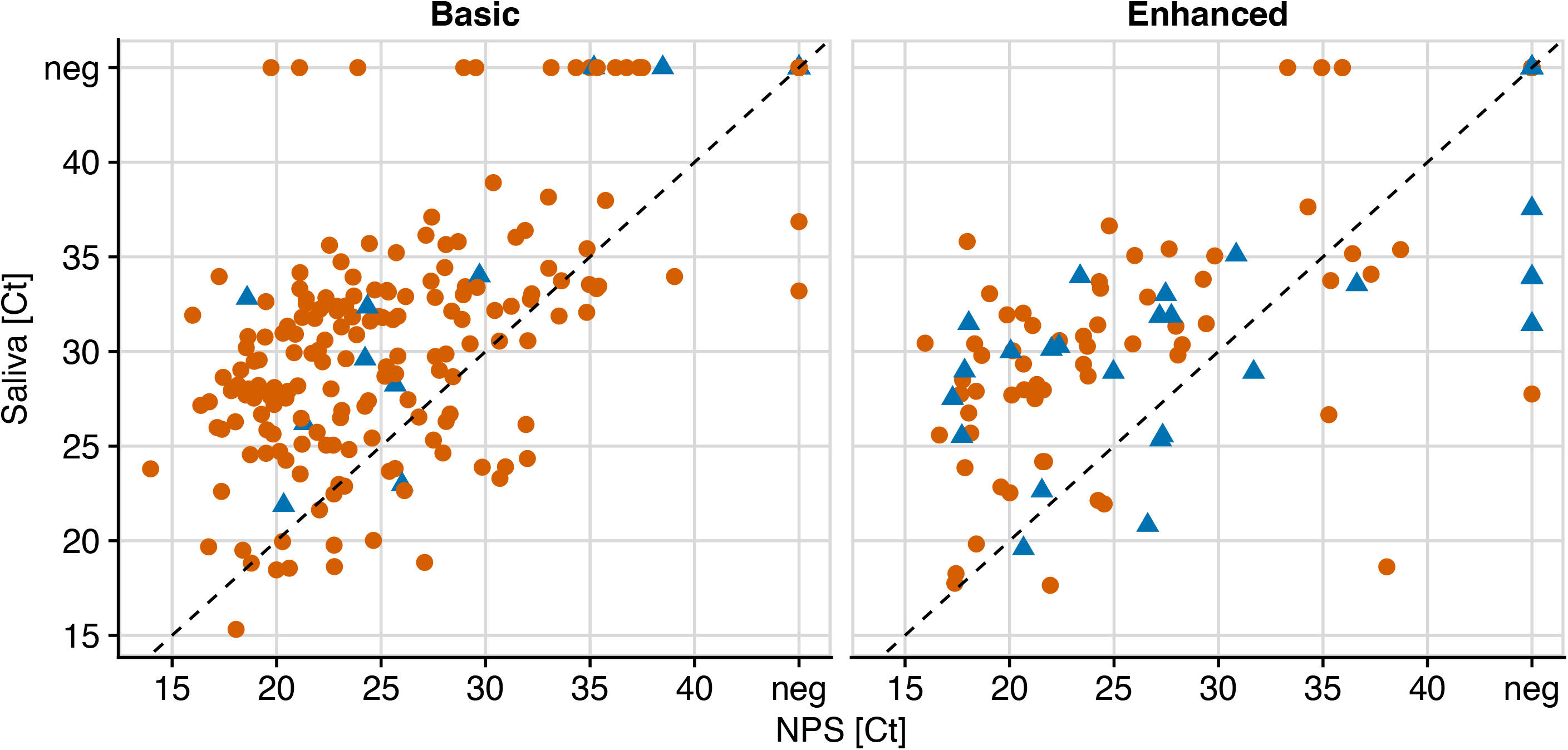
Intensified saliva sampling increases low level SARS-COV-2 detection in saliva. E-gene Ct values of paired NPS and saliva samples of study arm “Basic” (1-2x saliva per tube; N = 835) and “Enhanced” saliva collection (3x saliva per tube; N = 435). Red dots = adults, blue triangles = children.

Based on these findings, local health authorities in Zurich, Switzerland have launched mass testing for outbreak management utilizing the enhanced saliva sampling procedure in schools. We thus far conducted mass tests in 6 schools (N = 350-700 tests per school) and saliva sampling proved well accepted by children, parents and teachers, and easy applicable for mass collection and laboratory processing.

## Discussion

In the present study we sought to devise and evaluate a saliva sampling strategy that provides i) representative sampling of virus containing material, ii) easy and safe collection, iii) comfort for repetitive testing in adults and children, iv) possibility for home collection, v) straight forward processing in the laboratory. As a key element, saliva collection should not create infectious waste and should not need supervision by medical personnel to make it amenable for safe collection at home or in public institutions, for instance schools. We opted for a saliva collection procedure where participants clear their throat to first generate saliva from the back of the throat and then expectorate the saliva into an empty container. We considered clearing the throat important to sample material from the posterior oropharynx where SARS-CoV-2 sampling by oropharyngeal swabs is known to be efficient [34, 35]. While gargling with saline or buffer solutions has been suggested as a possibility to sample saliva from the deep throat [36, 37], we rated this procedure as less operable as the gargling solution would need to be optimized for taste to be accepted by individuals, could not include preservatives, and gargling itself may potentially generate aerosols. In addition, gargling is not practicable for many younger children, for whom we particularly sought to create more possibilities for SARS-CoV-2 testing, as NPS collection is often not practical in children. Our study demonstrates an excellent agreement of saliva in the head-to-head comparison with NPS and thus recommends saliva as alternate material for SARS-CoV-2 detection by RT-PCR. Up to a Ct 33 (equivalent to approximately 26’000 genome copies/ml) in the corresponding NPS, a notably high PPA (97.8% for the full cohort) is reached. Of note, virus loads in an even lower range are considered to impose a marginal risk for transmission as suggested by contact tracing and in vitro culturing studies [38-40]. Saliva performed equally well in children (PPA = 93.3%), notably, more saliva tested positive compared to NPS in this population (PPV = 84.8%). The reduced efficacy of NPS in children underlines the difficulty in obtaining correct swabs from them, highlighting the potential of saliva collection particularly when diagnosing children.

Considering the observed PPAs in detection, saliva may safely be envisaged as substitute for NPS detection in a range of settings. Possible scenarios include i) sampling of children, ii) home collection, iii) test centers without trained medical personnel (e.g. schools, universities, companies), iv) non-irritating alternative for persons that need frequent testing due to their occupation or health status, v) repetitive mass-testing. In situations where besides SARS-CoV-2 other respiratory viruses, e.g., Influenza and RSV, need to be excluded, NPS should, however, remain the standard material of choice as it allows rapid detection with multiplex-PCR from a single specimen. In addition, if SARS-CoV-2 infection has to be ruled out with the highest possible sensitivity (e.g. in transplantation), NPS should equally continue to be used.

The majority of SARS-CoV-2 in saliva represents likely virus secreted from infected cells in the nasopharynx and is not locally produced. Collecting material from the posterior oropharynx may thus be important. This is also highlighted in our study as the collection protocol with intensified throat clearing shows a trend to increased PPA at low viral loads. It remains possible that eating or drinking shortly before collection may decrease viral content in the oral cavity and throat. In the present study, neither eating, drinking nor smoking was controlled as study subjects came for an elective analysis by NPS and thus could only be informed about the saliva sampling on site immediately before the collection. Abstaining from food and beverage uptake shortly (1h) before saliva collection could be considered in forth-coming applications of saliva as test material, as it may increase the efficacy of SARS-CoV-2 detection in saliva even further.

In summary, our analysis rates saliva as valid alternate specimen for SARS-CoV-2 detection by RT-PCR, which is even superior in children when compared to NPS. Saliva collection is non-invasive, thus not strenuous for patients, does not need trained personnel, allows collection at any location, and allows self-collection. Importantly, as we show here, saliva collection does not require any adjustments in the diagnostics tests; established RT-qPCR can be used. Of note, while using RT-PCR as method for saliva testing causes only a minor loss in sensitivity compared to NPS, loss in sensitivity of rapid antigen tests needs to be carefully weighed when saliva is used as test material, as sensitivity of antigen tests is lower already in NPS. Saliva-RT-PCR testing has a further advantage over screens based on rapid antigen tests, as RT-PCR collected material allows for immediate subsequent analysis of positives as currently necessary for variant tracking by PCR and surveillance sequencing. Combined with the high reliability in detecting SARS-CoV-2 infection as demonstrated in our head-to-head comparison with the standard NPS, increasing and facilitating test efforts by monitoring SARS-CoV-2 infection in saliva is rapidly attainable. Saliva combined with RT-PCR particularly should be considered to improve testing opportunities in children as means to circumvent school closing.

## Data Availability

All data referred to in the manuscript are available from the corresponding author upon request.

## Acknowledgments

We thank Martin Ringer and the staff of the participating test centers for coordinating the sample collection, the staff of the Institute of Medical Virology diagnostics unit, sample triage and administration for their support and Urs Karrer and Alexander Wepf for helpful discussions.

